# Perceptions and Attitudes Towards Shoulder Padding and Shoulder Injury in Rugby Union

**DOI:** 10.1101/2020.05.15.20102905

**Authors:** Angus Hughes, Prof. Matt Carré, Dr. Heather Driscoll

## Abstract

**Objective:** To develop an understanding of the role of shoulder padding in preventing injuries in rugby by investigating player perceptions and attitudes towards shoulder padding and extending previous research into the nature of shoulder injuries in rugby.

**Methods:** A survey was distributed to current rugby players over 13 years old. Questions related to the participants’ demographic, attitudes to shoulder padding and shoulder injury history.

**Results:** 616 rugby players responded to the survey. 66.1% of respondents had worn shoulder padding at some point during their career. Youth players (13–17 years old) and the older demographic (36+ years old) perceived shoulder padding to be more effective. 37.1% of respondents considered shoulder padding to be effective at preventing cuts and abrasions with 21.9% finding it very effective. 50.3% considered it to be effective at preventing contusion injury with 9.7% finding it very effective. 45.5% wore padding for injury prevention, while 19.2% wore padding to protect from reoccurring injury. 38.6% did not wear shoulder padding because they felt it was not needed for the game of rugby. Sprain/ ligament damage (57.5%) and bruising (55.5%) to the shoulder were the most commonly reported injury.

**Conclusions:** Research should focus on quantifying the injury preventive capabilities while also educating the rugby community on shoulder padding. Bruising, cuts and abrasion injuries to the shoulder are prevalent. The ability of shoulder padding to protect from these injuries should be further explored.

## BACKGROUND

Rugby union is a collision sport, resulting in a relatively high injury rate (90.1 per 1000 player match hours (PMH)), when compared with soccer (64.4 per 1000 PMH) and tennis (31.1 per 1000 PMH)^1^. On average, one rugby match leads to 456.8 impacts^2^, these impacts are mostly seen in the tackle (48%). 65% of shoulder injuries are caused in the tackle^3^, therefore, although not as prevalent as lower limb injuries (50.6 per 1,000 PMH^4^) shoulder injuries have a substantial incidence rate in rugby union (12.7 per 1,000 PMH^5^). The epidemiology of shoulder injury in rugby union has been reported^5,6^. However, the definition of injury used in this research (24+ hour time loss from all participation) creates suspicion that less severe injuries including bruising, cuts and abrasions are under reported.

Shoulder padding is a popular addition to most rugby players’ equipment. It possesses properties that allow it to dissipate a certain amount of impact energy resulting in it being reported that 70% of players will wear shoulder padding to reduce the risk of injury^7^. However, the ability of shoulder padding to reduce injury has not been quantitatively assessed, therefore should not be considered as a means of injury prevention. Coupled with this, regulations have been set by rugby’s governing body World Rugby to limit its impact protection potential, and as such World Rugby do not view shoulder padding as a form of significant protection. It is however, considered to reduce the risk of superficial injuries like lacerations as well as add a certain amount of comfort to the impacts seen in rugby.

Padded equipment including shoulder padding and headgear in rugby union is designed with a focus on its ability to dissipate impact forces and support vulnerable body structures. Recent research has explored the use of protective headwear in rugby union with 67% of rugby players having worn this form of padded equipment^8^. However, players’ attitudes towards the use of shoulder padding is generally unresearched. These attitudes can have an influential effect on the use of shoulder padding. Knowledge of players’ behaviors towards shoulder padding should be established in order to understand the role of shoulder padding in injury prevention and to help develop new products and methodologies with which to assess their performance.

The current study therefore seeks to develop an understanding of player perceptions and attitudes towards shoulder padding as well as extend previous research regarding shoulder injury in rugby. First, the study aims to develop detailed knowledge of players’ attitudes and perceptions of shoulder padding through a mixed methods design, while examining how different sub groups may differ in their perceptions and attitudes. Secondly, the study aims to examine shoulder injury epidemiology of rugby players, including any effects of players’ attitudes and perceptions of shoulder padding.

## METHODS

### Survey Development

An online survey was developed to investigate the attitudes and perceptions of shoulder padding as well as the shoulder injury history of rugby players. During the preparation of this study, 25 rugby players contributed to the development of the survey through commenting on an initial set of pilot questions. After evaluation of this pilot, a final questionnaire was presented as an online survey using the software Google Forms.

Section 1 of the survey collected demographic information including gender, age, playing experience, level and position. Section 2 then collected participants’ attitudes and perceptions to shoulder padding and included questions regarding shoulder padding usage, reasons for wearing and not wearing shoulder padding using open ended text box style questions, as well as participants’ perceptions of how effective shoulder padding is with regards to injury prevention both generally using a 5-point Likert scale (1=‘not at all’, 5=‘a great deal’) and specifically to certain injuries using a different 5-point Likert scale (1=’very ineffective’, 5=’very effective’). Section 3 then collected information regarding the participants’ shoulder injury history so that shoulder pad usage and attitudes could be linked with shoulder injury experience as well as add to epidemiological data. The questionnaire included both closed and open questions. This mixed methods design allowed for descriptive and interpretive information to be obtained.

### Survey Deployment

Rugby players older than aged 13 of any gender and skill level were targeted during the deployment of the questionnaire. The questionnaire was distributed to respondents between May and July 2018. The questionnaire was publicised through various social media platforms including directly through Word Rugby’s twitter handle. Various rugby clubs were also approached, and the survey link was sent to its members. The country in which the respondents resided was not controlled.

### Data Analysis

Quantitative data was inputted into SPSS (version 25) and descriptive statistics were produced in order to examine demographics, shoulder pad usage, and shoulder injury history. One-way Anova analysis was performed to compare mean differences between perceived effectiveness of padding and demographic information. Open ended survey responses (reasons for wearing and not wearing shoulder padding) were examined using a thematic approach, as used by Braun and Clark^9^. Eight higher order themes were identified for the open ended questions. Descriptive statistics for these themes were then produced in order to examine the responses.

## RESULTS

### Basic Characteristics

At total of 616 responses were collected from the survey, giving a wide demographic of rugby players (Table 1).

**Table 1.**
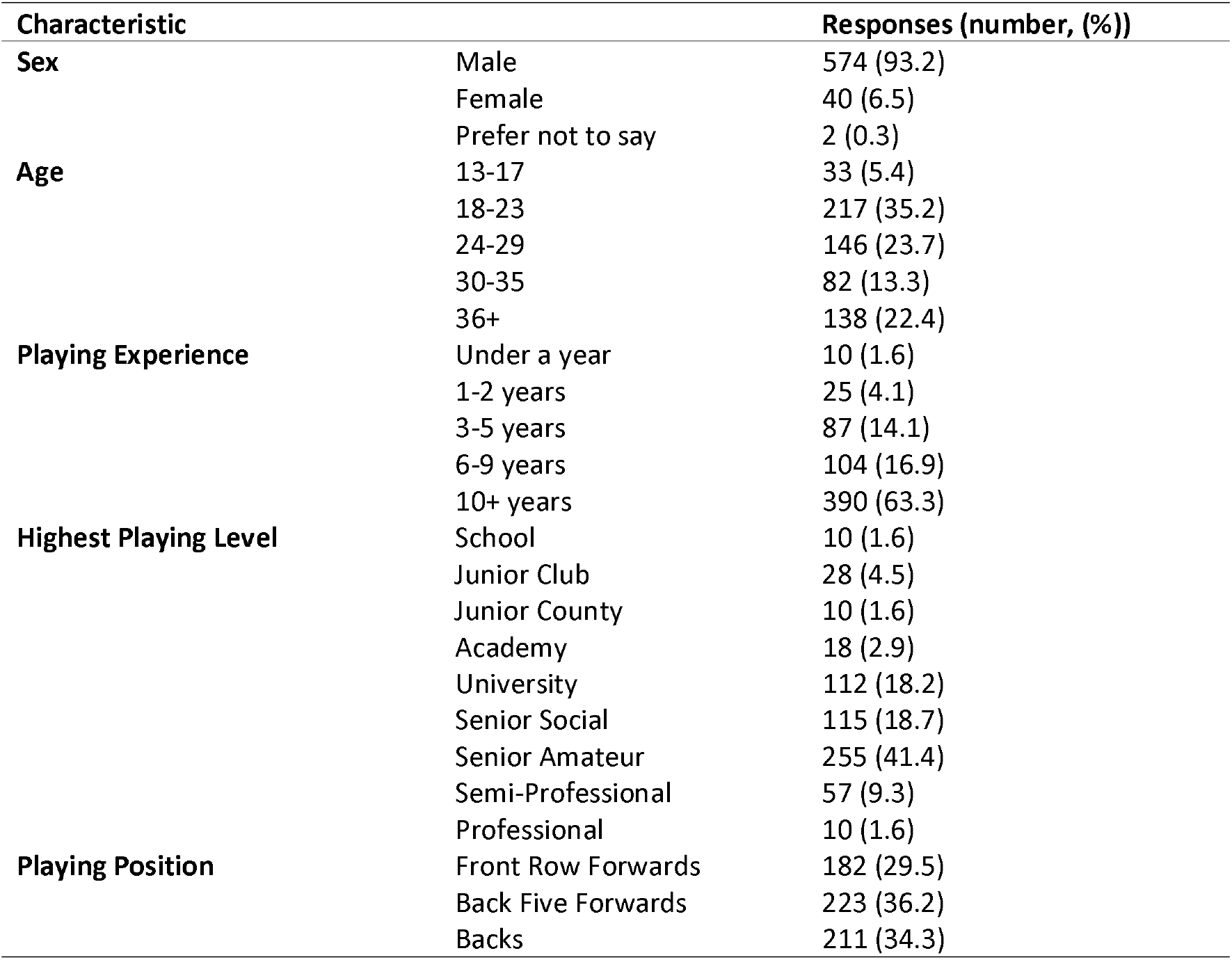
Demographic information of players surveyed.

### Shoulder pad use

Of all the players who completed the questionnaire (n=616), 66.1% (n=407) had worn shoulder padding at some point. 33.9% (n=209) had never worn shoulder padding. From those who had worn shoulder padding, 9.9% (n=61) always wore shoulder padding, 17.7% (n=109) only wore shoulder padding during matches, 13.1% (n=81) wore shoulder padding, but only because of an injury and 25.3% (n=156) wore shoulder padding regularly in the past but at present did not. 61% (n=111) of front row forwards had worn shoulder padding at some point, 61% (n=136) of back five forwards had worn shoulder padding at some point while 74% (n=129) of backs had worn shoulder padding at some point.

### Attitudes towards effectiveness of shoulder padding

Of the players who completed the questionnaire, the mean perception (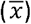) of the effectiveness (Likert scale 1–5) of padding was 2.44. A mid-level response would be 3. When player’s shoulder padding usage behaviours were factored in, the results were as follows, those that always wore shoulder padding (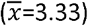), only wore shoulder padding in matches (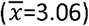), wore shoulder padding, but only because of an injury (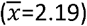), wore shoulder padding regularly in the past but at present did not (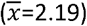) and had never wore shoulder padding (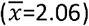). It was found that gender (p=0.305), playing position (p=0.161) and playing level (p=0.513) had no significant (p > 0.05) association with the perceived effectiveness of padding. However, age (p=0.003) and playing experience (p=0.008) did (p < 0.05). The age group 24–29 found padding least effective (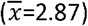) with the 13–17 age group finding it most effective (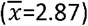). Respondents with the most playing experience (10+) found padding least effective (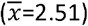), while respondents with 1–2 years of experience found padding most effective (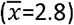).

When considering specific injuries, the perceived effectiveness of shoulder padding is seen in figure 2. 37.1% of respondents considered shoulder padding to be effective at preventing cuts and abrasions with 21.9% finding it very effective. 50.3% considered it to be effective at preventing contusion (bruising) injury with 9.7% finding it very effective. Perceived effectiveness of shoulder padding then drops with more severe injuries, 17.4% of respondents considered it either effective or very effective at preventing sprain/ ligament damage, as well as 10.6% for dislocation and 21.5% for bone injury.

**Figure 2:**
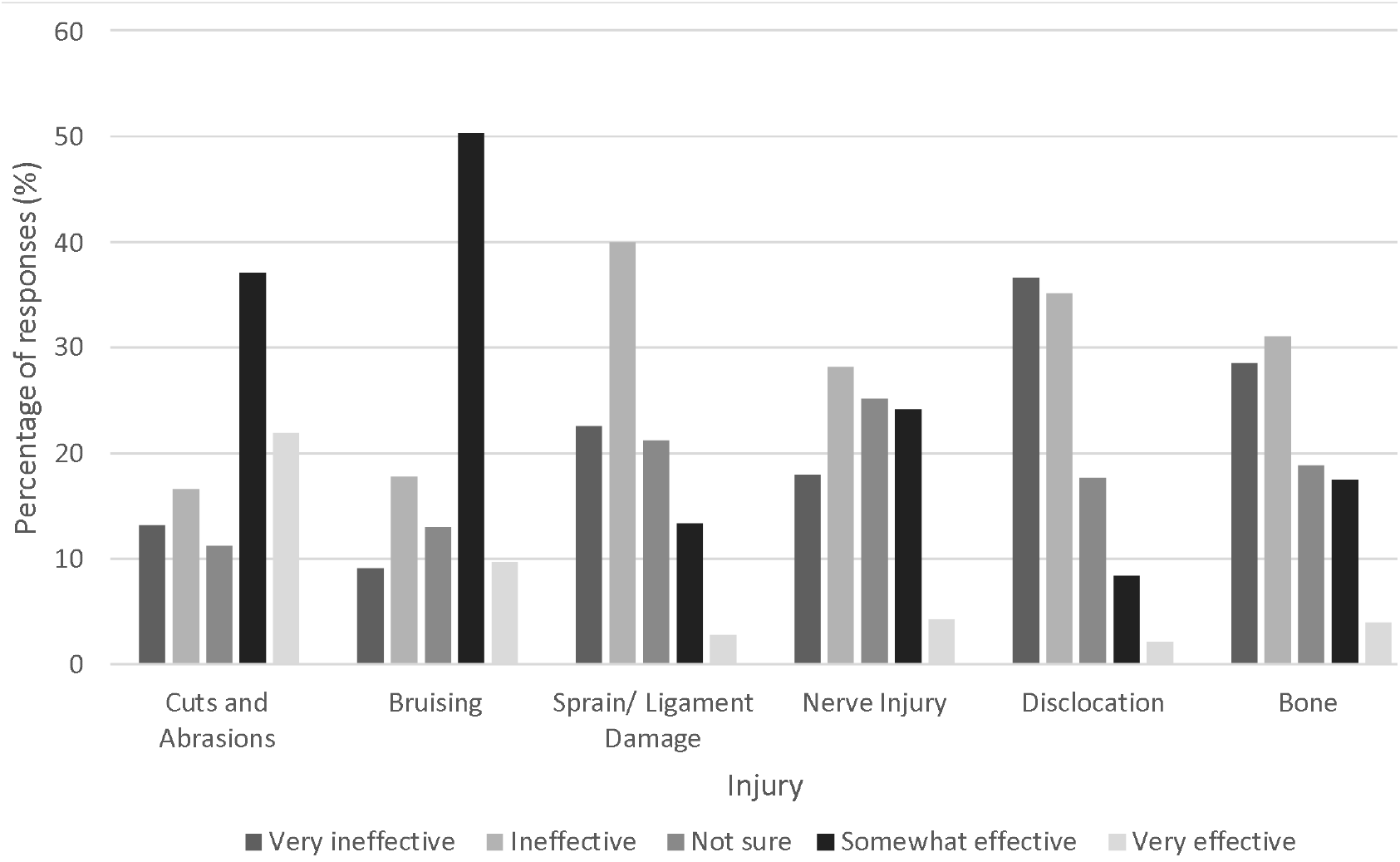
The perceived effectiveness of padding for specific injuries

### Attitudes of players who wear shoulder padding

After the responses were analysed, eight themes emerged when considering the reasons why players wore shoulder padding (table 2). Respondents first response was taken and there were 386 responses in all. 62.6% of responses indicated wearing shoulder padding as a form of protection or injury prevention with 19.2% of these being to protect from a reoccurring injury. 15.8% of responses implied rugby players wore shoulder padding to feel more confident, mainly in the tackle situation. 9.3% of responses indicated wearing shoulder padding for comfort in impacts rather than as a form of protection.

**Table 2.** Reason themes for wearing shoulder padding (listed from most to least common)

| **Higher order themes (n=386)** | **Example Responses** |
| --- | --- |
| **Injury Prevention and Padding (43.5%)** | Protection.<br>Protect from minor shoulder injury.<br>Degree of protection offered to shoulder and collar bone in contact.<br>Protect against soft tissue injury. |
| **Protection from reoccurring injury (19.2%)** | To protect my shoulder whilst it wasn't 100%.<br>Returning from an injured shoulder.<br>To reduce impact on shoulders following an injury.<br>Damaged my ac joint and padding it was the only way I could tackle with the least amount of discomfort. |
| **Confidence (15.8%)** | When I first played contact rugby, it gave me greater confidence when making a tackle.<br>Confidence in the tackle area.<br>Purely confidence. I don't believe it helps, other than my mind.<br>Feel more secure.<br>It makes me feel more confident about making tackles in matches. |
| **Comfort in impacts (9.3%)** | Just gives a little bit of extra comfort in the pack for tacking and scrums.<br>Less sore shoulders after scrum.<br>To stop my shoulders being rubbed raw in scrums. |
| **Recommendation from coaches, friends or parents (7.3%)** | When I was younger I wore it for shoulder protection mainly on the insistence of my Mum.<br>Was recommended by the coach.<br>It was popular to wear them. |
| **Habit (1.8%)** | It feels part of my gear, same as gumshield, shorts etc.<br>Was given to me for free, got used to wearing it and then didn't like the feel of playing without it. |
| **To change own physical appearance (1.6%)** | Being smaller than everyone else.<br>Due to my size frame shoulder pads helped make me feel bigger, it had a bit of placebo effect. |
| **To try it out (1.6%)** | No specific reason, a friend gave it to me and I decided to try it out. |

### Attitudes of players who do not wear shoulder padding

Eight themes were identified when considering players who did not wear shoulder padding (table 3), Respondents first response was taken and there were 352 responses in all. 38.6% of responses indicated wearing shoulder padding was not needed, with 21.3% of responses indicating shoulder padding was uncomfortable. 16.8% of responses indicated rugby players did not feel padding had added protective benefits.

**Table 3.** Reasons for not wearing shoulder padding.

| Higher order themes (n=352) | Example Responses |
| --- | --- |
| **They are not required (38.6%)** | I stopped wearing it as I didn't need them to absorb impacts anymore. Just never bothered with it.<br>I don't see the need for shoulder padding, I've never hurt my shoulders before.<br>Injury healed so no longer required shoulder pad protection. |
| **Discomfort (21.3%)** | I stopped as it was uncomfortable and I tended to overheat.<br>Can get too hot wearing them and sometimes uncomfortable.<br>I get too hot wearing them otherwise I would probably wear them all the time.<br>I feel claustrophobic in them at times and get too hot. |
| **Do not offer protection (16.8%)** | I am unaware of the difference it could make to my safety or skills.<br>Didn't seem to help with anything as so thin.<br>No added benefits to protection. |
| **Restricts movement (6.3%)** | It adds bulk, makes it harder to manoeuvre.<br>Movement limiting.<br>My movement felt restricted with the pads, and I wanted full movement to avoid injury. |
| **Cost and Availability (6.3%)** | It seems unnecessary and is an expense I can't really afford.<br>Too costly to replace. |
| **Impacts the game negatively (4%)** | I enjoy the hard-hitting nature of the game which I feel would lack with pads.<br>Not wearing shoulder padding encourages a correct technique in tackle/contact situations and observation of the laws of the game.<br>Wearing padding too easily encourages reckless and undisciplined hits from bad angles with greater force.<br>Enjoying the tackle more without them. |
| **Stigma (3.7%)** | Not the manly thing to do.<br>It's for girls.<br>There is a perception of people who wear padding being 'soft'. |
| **False sense of security (3.1%)** | It gives a false sense of security, if you're going to break your bones, you're going to break your bones.<br>Disagree with it. I believe it gave a false belief to those who did. |

### Shoulder injury data

Of the players who completed the questionnaire, 72.8% (n=447) reported having a shoulder related injury as a result of playing. 27.2% (n=167) had not had a shoulder related injury as a result of playing. Of those that reported having a shoulder related injury as a result of rugby, 35.8% (n=160) reported experiencing a cut or abrasion injury, 55.5% (n=248) reported experiencing a contusion injury, 57.5% (n=257) reported experiencing a sprain/ ligament related injury, 33.1% (n=148) reported experiencing a nerve related injury, 18.1% (n=81) experienced a dislocation and 20.0% (n=89) experienced a bone related injury. Figure 2 displays specific shoulder injury history as a function of shoulder padding usage. Backs sustained less shoulder injuries (66%), when compared to front row forwards (79%) and back five forwards (74%). When comparing shoulder injury data between players that always wear shoulder padding and players that have never worn shoulder padding, more back five forwards that always wore padding had sustained a shoulder injury (76%) than those who had never worn it (60%). Similar results were seen with front row forwards, 89% of the front row that always wore padding had sustained an injury compared with the 66% that had never worn padding. However, 50% of the backs that always wore shoulder padding had sustained a shoulder injury, this was the same for the backs that never wore padding (50%).

**Figure 2.**
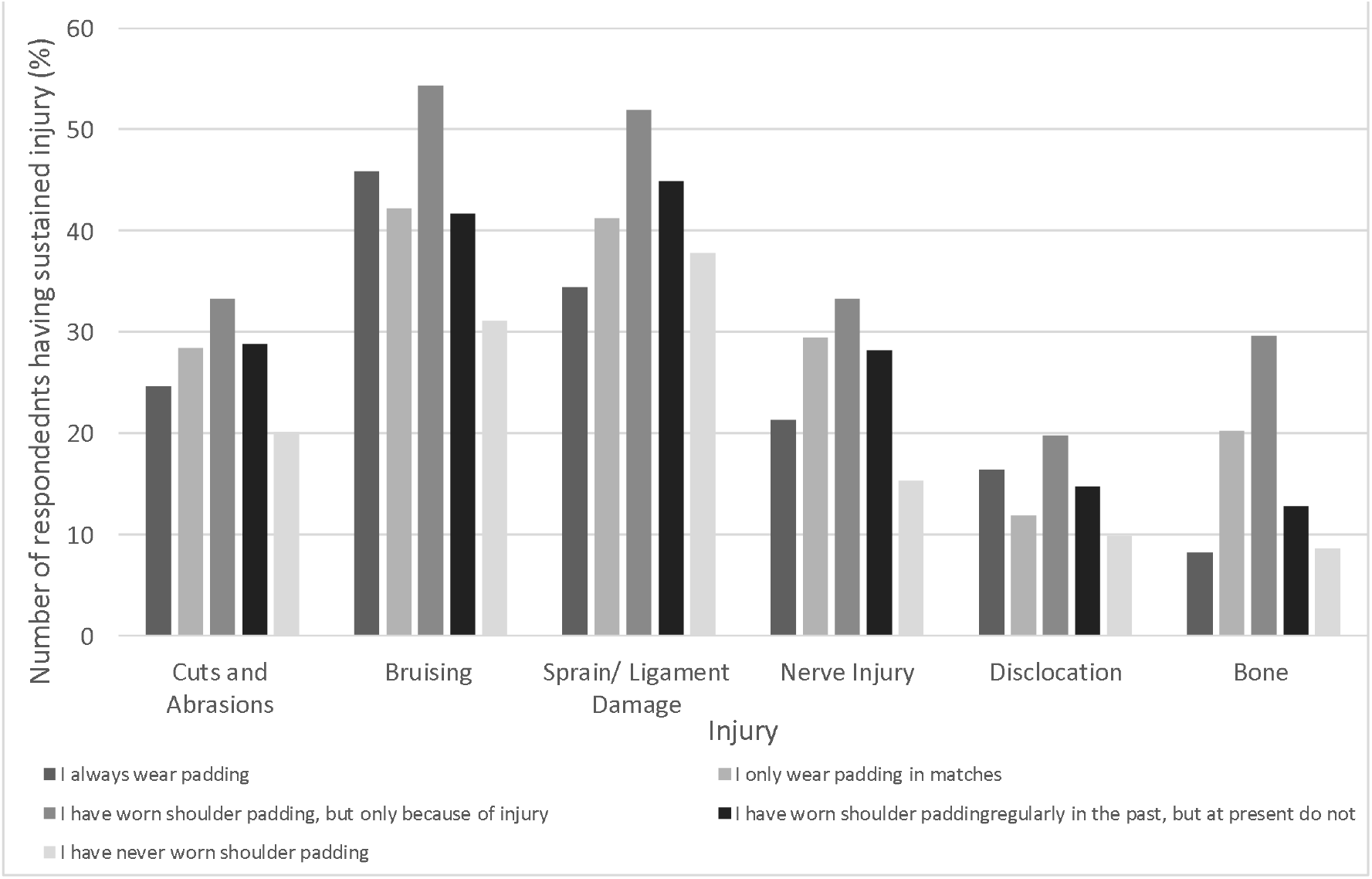
Specific shoulder injury history as a function of shoulder pad usage.

## DISCUSSION

The major findings of this study were that; (1) 66.1% of rugby players had used shoulder padding at some point in their playing career; (2) the primary reason for wearing shoulder padding was either as a means of injury prevention (43.5%) or to protect from reoccurring injury (19.2%); (3) the primary reason for not wearing shoulder padding was that it was not needed for the game of rugby(38.6%); (4) Sprain/ ligament damage (57.5%) and bruising (55.5%) were the most prevalent shoulder injuries, while (35.8%) of respondents had sustained a cut or abrasion to the shoulder, underreported in previous research.

## Shoulder Padding

Results indicated that mean perceived effectiveness of padding increased with increased use. Mean effectiveness of padding was 3.33 (Likert scale 1–5) for players who always wore shoulder padding but 2.19 for players who have but do not currently wear padding. Age influenced the perceived effectiveness of padding with youth (13–17 years old) as well as older (30–35, 36+ years old) respondents having an increased belief on the effectiveness of padding at preventing injury. This is possibly due to the lower collision forces experienced at youth level while still wearing the same thickness of padding. Both a youth and an aged population may also see padding as more effective due to the fact they are more susceptible to injury in rugby^10^. Whilst there seems to be a good awareness into the limitations shoulder padding has at preventing injury, further education should be directed at youth level in order to reinforce player knowledge.

59% of respondents considered shoulder padding to be either effective or very effective at preventing cuts and abrasions with 60% of respondents considering shoulder padding to be either effective or very effective at preventing contusions. These results compliment previous research into padded headgear, finding 55% of respondents to consider headgear to be effective at preventing minor injuries^8^. Clearly, shoulder paddings ability to reduce the risk of superficial injuries like cuts and bruising must be measured in order to justify rugby players’ perceptions of padding. 10.6% considered shoulder padding to be either effective or very effective at preventing dislocations, as well as 21.5% considering shoulder padding to be either effective or very effective at preventing bone injury. Although these percentages are low, further education, as well as other injury preventative methods like the BokSmart^11^ initiative in South Africa which coaches correct tackle technique should be considered to ensure fewer rugby players view shoulder padding as an effective tool at preventing severe injuries to the shoulder.

The primary reasons for wearing shoulder padding were either as a means of injury prevention (43.5%) or to protect from reoccurring injury (19.2%). This was to be expected due to how shoulder padding is commercially branded as well as it proven impact attenuating abilities^12^. 15.8% of players wore shoulder padding to increase confidence, mainly in the tackle. The outweighing association between shoulder pad use and injury prevention suggests this increased confidence stems from a decreased worry about getting injured. This result is similar to a study by Barnes et al.^8^. 13% of responses related to increased confidence as a motivation for its use. There is a clear link that wearing any form of padding in rugby may lead to an increased confidence to not get injured. It is however, important to note World Rugby does not view shoulder padding as a form of protective equipment and has set impact attenuating abilities to a maximum limit, changes in player behaviours can therefore be limited. Contrary to this, it has been suggested that some players can become overly reckless when wearing protective equipment^13^. This is further backed up by 3.1% of reasons for not wearing shoulder padding being related to the feeling of a false sense of security. These factors could explain why an increased percentage of players who always wear shoulder padding had sustained a shoulder injury as a result of playing rugby than those that had never worn shoulder padding.

The primary reason for not wearing padding was that shoulder pads were not needed in rugby (38.6%). Previous research suggests the physical nature of the game leads to players adopting a mind-set where extra padding is not needed^14^. Discomfort (21.3%) and the feeling of restricted movement (6.3%) were also key reasons for not wearing padding. Similar to research into padded headgear in rugby, which also found discomfort and heat regulation issues to be primary reasons for not wearing padded headgear^15^. 16.8% of respondents felt shoulder padding offered no extra protection. Further research into what injuries shoulder padding may reduce the risk of is needed followed by education of these findings to rugby players. Manufacturers should consider the factors of discomfort and restricted movement while also acknowledging World Rugby regulations when designing future products.

## Shoulder Injury

When asked what specific shoulder injuries the participants had sustained, sprain/ ligament damage (57.5%) and contusion injuries (55.5%) were the most prevalent. Previous research reports a lower frequency of contusion injuries (12 – 17%^3,5^). Possibly due to the injury definition used in both studies which would lead to the underreporting of a contusion that may not be of the severity to cause time loss or require medical attention. As well as this, it is possible players were more likely to respond to the survey if they had had a shoulder injury. The large prevalence of reported contusion injuries to the shoulder does however suggest shoulder paddings ability to decrease the risk of a contusion should be explored. This also the case with cuts and abrasion injuries, 35.8% of respondents had sustained a cut or abrasion as a result of playing rugby. No published research reports cuts, lacerations or abrasions specifically to the shoulder region.

When comparing the occurrence of specific injuries with the participants’ use of padding, more players that always wore padding (70%) had sustained a shoulder injury than those that that had never worn padding (60%), possibly because of the suggestion that wearing padding can increase the risk of injury due to an increased tendency to use reckless poor tackle technique. With regards to less severe injuries, players that always wore padding had sustained more cuts and abrasions (24.6%) as well as contusion injuries (45.9%) than that of players that had never worn padding (20.1%, 31.1%). Generally, players that had never worn padding felt they did not see the need to wear it, with the primary reason for wearing shoulder padding being as a means of protection, this group of participants’ may not have needed the added protection of shoulder padding, therefore explaining the larger reporting of less severe injuries in players that always wear padding. Coupled with this, some players that had never worn padding did so out of stigma. The stigma of wearing padding may also have led to the under reporting of less severe injuries like cuts, abrasions and bruising.

## Limitations

Limitations of this study arise from the method of data collection, recall bias^16^ may have been an issue due to the self-reporting style of data collection. However, the varied demographic of respondents would have greatly reduced selection bias. Self-reporting of previous injuries may also have been an issue; future studies should look to use injury data that has been reported by medical professionals. The study was the first to explore the attitudes of rugby players to shoulder padding specifically, while also linking shoulder pad use with shoulder injury history. Future studies should explore whether shoulder pad use affects actual playing behavior as well as shoulder injury occurrence.

## Conclusions

To conclude, the primary reason for wearing shoulder padding was as a means of injury prevention or protection. However further education is needed so that players are aware of the protective limitations shoulder padding entails. The study presents new findings that less severe shoulder injuries have been underreported in previous research due to the injury definition being used. The ability of shoulder padding to reduce the risk of these less severe injuries should be quantified while also educating rugby players that shoulder padding will not prevent injury and may encourage overly reckless poor tackle technique. The findings from this can be used to facilitate the development of new products and methodologies with which to assess their performance.

## Data Availability

Raw data available on request from the author

## What are the new findings?

- 66.1% of rugby players had used shoulder padding at some point in their playing career.
- The primary reasons for wearing shoulder padding was either as a means of injury prevention (43.5%) or to protect from reoccurring injury (19.2%).
- The primary reasons for not wearing shoulder padding was that it was not needed for the game of rugby (38.6%) or was perceived to be uncomfortable (21.3%).
- Underreported in previous research, players sustain significant bruising (55.5%) as well as cuts and abrasions (35.8%) to the shoulder while playing rugby union.

## How might it impact on clinical practice in the future?

- Education and research into the injury protective capabilities of padded clothing in rugby should be addressed.

## Acknowledgments

This project is funded by World Rugby and the Engineering and Physical Sciences Research Council (EPSRC).

## Ethical Approval

The University of Sheffield ethics board gave ethical approval prior to the project, participants gave informed consent before completing the survey.

## Data Deposition

Due to the nature of this research, participants of this study did not agree for their data to be shared publicly, so supporting data is not available

